# SymScore: Machine Learning Accuracy Meets Transparency in a Symbolic Regression-Based Clinical Score Generator

**DOI:** 10.1101/2024.10.28.24316164

**Authors:** Olive R. Cawiding, Sieun Lee, Hyeontae Jo, Sungmoon Kim, Sooyeon Suh, Eun Yeon Joo, Seockhoon Chung, Jae Kyoung Kim

**Affiliations:** Department of Mathematical Sciences, KAIST, Daejeon, 34141, Republic of Korea; Biomedical Mathematics Group, Pioneer Research Center for Mathematical and Computational Sciences, Institute for Basic Science, Daejeon, 34126, Republic of Korea; Department of Mathematics, Kyungpook National University, Daegu, 41566, Republic of Korea; Division of Applied Mathematical Sciences, Korea University, Sejong, 30019, Republic of Korea; Department of Psychology, Sungshin Women’s University, Seoul, 02844, Republic of Korea; Department of Neurology, Neuroscience Center, Samsung Medical Center, Sungkyunkwan University School of Medicine, Seoul, 06351, Republic of Korea; Department of Psychiatry, Asan Medical Center, University of Ulsan College of Medicine, Seoul, 05505, Republic of Korea

**Keywords:** Medical questionnaires, Interpretable machine learning, Shortened questionnaires, Symbolic regression, Risk score evaluation, Clinical decision making, Explainable artificial intelligence

## Abstract

Self-report questionnaires play a crucial role in healthcare for assessing disease risks, yet their extensive length can be burdensome for respondents, potentially compromising data quality. To address this, machine learning-based shortened questionnaires have been developed. While these questionnaires possess high levels of accuracy, their practical use in clinical settings is hindered by a lack of transparency and the need for specialized machine learning expertise. This makes their integration into clinical workflows challenging and also decreases trust among healthcare professionals who prefer interpretable tools for decision-making. To preserve both predictive accuracy and interpretability, this study introduces the Symbolic Regression-Based Clinical Score Generator (SymScore). SymScore produces score tables for shortened questionnaires, which enable clinicians to estimate the results that reflect those of the original questionnaires. SymScore generates the score tables by optimally grouping responses, assigning weights based on predictive importance, imposing necessary constraints, and fitting models via symbolic regression. We compared SymScore’s performance with the machine learning-based shortened questionnaires MCQI-6 (*n* = 310) and SLEEPS (*n* = 4257), both renowned for their high accuracy in assessing sleep disorders. SymScore’s questionnaire demonstrated comparable performance (MAE = 10.73, *R*^2^ = 0.77) to that of the MCQI-6 (MAE = 9.94, *R*^2^ = 0.82) and achieved AU-ROC values of 0.85-0.91 for various sleep disorders, closely matching those of SLEEPS (0.88-0.94). By generating accurate and interpretable score tables, SymScore ensures that healthcare professionals can easily explain and trust its results without specialized machine learning knowledge. Thus, SymScore advances explainable AI for healthcare by offering a user-friendly and resource-efficient alternative to machine learning-based questionnaires, supporting improved patient outcomes and workflow efficiency.

## 1. Introduction

Recent advancements in artificial intelligence (AI) have revolutionized healthcare, impacting disease diagnosis, drug discovery, patient management, and personalized treatment plans [1, 2, 3, 4, 5, 6]. In particular, sentiment analysis and other machine learning methods have shown promise in understanding patient data and improving predictive accuracy [7, 8, 9]. However, despite these benefits, the complexity and lack of transparency in AI systems, along with real-world failures of AI-driven healthcare tools, have limited their practical application in medical settings, leading to skepticism and hesitation among healthcare professionals [10, 11]. For example, an external validation of Epic System’s sepsis prediction algorithm revealed that the model produced many false alarms, which resulted in clinicians ignoring the system’s recommendations [12]. Furthermore, studies examining the views of clinicians regarding the adoption of AI-driven medical systems show that many, while recognizing the advantages of AI, have reservations regarding their clinical applicability [13, 14, 15]. Thus, despite AI’s potential to streamline diagnosis, many clinicians prefer to rely on their own expertise and manual assessments. Common themes among their reservations include a lack of explainability and poor integration of AI tools into existing clinical workflows, which can render these tools more burdensome than helpful [16, 17]. These challenges led to the emergence of the field of explainable AI (XAI), focusing on creating transparent and understandable AI systems, thereby enhancing trust—a crucial factor in healthcare [18, 19].

One key aspect of XAI is the development of interpretable machine learningbased models that can be easily understood and trusted by healthcare professionals. Tools like SHAP (SHapley Additive exPlanations) [20] and the ELI5 (Explain Like I’m 5) [21, 22] have been instrumental in this regard. They provide explanations for AI model predictions by assigning importance values and weights to features. For instance, SHAP has been used to interpret models predicting cardiac health risks [23], hypertension [24], and sleep disorders [25], while ELI5 has helped explain AI-driven classifications of clinical depression [26] and stroke prediction models [27]. These tools strive to balance predictive accuracy with transparency, making them suitable for healthcare.

The need for interpretable and efficient self-report questionnaires in disease diagnosis and risk assessment is well-recognized [28, 29]. Traditionally, these questionnaires have been lengthy, which can lead to lower response rates and compromised data quality due to respondent fatigue [30]. In response, shorter versions have been developed using techniques such as principal component analysis and factor analysis [31, 32, 33, 34]. While they have been widely used, these methods have limitations, including subjective decisions and assumptions of normal data distribution, compromising predictive accuracy [35, 36, 37, 38, 39]. These assumptions often compromise predictive accuracy and limit the generalizability of the results, particularly in diverse clinical settings.

Machine learning-based shortened questionnaires have emerged as an alternative, offering accurate predictions with fewer questions [25, 30, 40, 41, 42, 43, 44, 45, 46, 47, 48, 49]. However, these models often face practical challenges in clinical settings [40, 50]. Integrating machine learning methods into existing medical systems can be complex and resource-intensive. Additionally, the application of these methods requires extensive training or the need for specialized professionals, both of which are costly and time-consuming. Moreover, these machine learning-based questionnaires are often seen as ‘black boxes,’ making them difficult for medical professionals to understand and trust. Although interpretable tools such as SHAP, LIME, and ELI5 provide feature importance, they do not offer the same level of transparency as the original questionnaires, which yield direct scores that healthcare professionals can interpret straightforwardly. This study is therefore motivated by the need to develop a clinical score generator that produces score tables for shortened versions of these questionnaires, allowing for the estimation of the results of the original questionnaires. Our approach ensures that the weights assigned to responses transparently reflect their contribution to disease severity while preserving the monotonic relationship of key items with disease severity. This approach builds trust with healthcare professionals and facilitates smoother integration into clinical workflows, without sacrificing accuracy or requiring specialized expertise.

To tackle transparency problems and align with XAI principles, Xie et al. [51] developed AutoScore, an automatic clinical score generator combining machine learning with regression modeling. AutoScore uses a random forest algorithm to select key questions from the original questionnaire and groups responses to form logistic models predicting risk scores. The simple conversion of coefficients of the models to the weights of responses leads to a user-friendly, shortened questionnaire. Thus, it has been widely adopted for various applications, such as predicting mortality in emergency patients [52], assessing acute kidney injury severity [53], evaluating Grave’s orbitopathy [54], and determining amyloid positivity [55]. Recently, La Cava et al. developed a symbolic regression-based tool called FEAT (Feature Engineering Automation Tool) [56], which automates feature construction and selection from high-dimensional electronic health record (EHR) data, providing simple and interpretable feature weights.

Despite their success, both AutoScore and FEAT have limitations, particularly in handling feature grouping, which can compromise accuracy. AutoScore, for instance, manually groups responses through trial and error, relying on subjective choices such as dividing age into quantiles. While FEAT automates this process, it only allows for two groups, limiting its flexibility. Additionally, neither method ensures that the assigned weights accurately reflect a monotonic relationship with disease severity. Lastly, both methods predict disease severity categories but do not predict the total score of the original questionnaire.

To overcome these limitations, we developed the Symbolic Regression-Based Clinical Score Generator (SymScore), which offers several key innovations not found in existing methods. SymScore leverages symbolic regression to automatically group item responses and determine optimal weights for each group. It incorporates essential constraints to ensure that response weights increase monotonically where appropriate, establishing a clinically meaningful relationship between responses and disease severity. This overcomes the manual and often subjective grouping of responses seen in AutoScore, and the limited flexibility in the grouping mechanism in FEAT. Moreover, SymScore supports both regression and classification tasks, which allows it to predict not only disease categories but also the total score of the original questionnaire. This capability is lacking in existing approaches and thus adding it provides flexibility for a wide range of clinical applications. By integrating these unique aspects of SymScore—automatic response grouping, the application of monotonicity constraints, and flexibility in both regression and classification, SymScore’s shortened questionnaires achieve accuracy comparable to that of existing machine learning-based questionnaires like MCQI-6 [43] and SLEEPS [25], which are specifically designed for predicting sleep disorders. This demonstrates that SymScore maintains transparency and ease of use without sacrificing accuracy, offering a straightforward alternative to complex machine learning algorithms. In conclusion, SymScore represents a significant advancement in XAI for healthcare, paving the way for widespread adoption of shortened, ready-to-use, and interpretable questionnaires. This approach fosters more efficient and informed clinical decision-making, ultimately contributing to enhanced patient outcomes and a more sustainable healthcare system.

### 1.1. Basic Concepts/ Preliminaries

#### 1.1.1. Machine learning-based shortened questionnaire, MCQI-6

The Metacognitions Questionnaire-Insomnia (MCQ-I) was designed to measure metacognitive beliefs about primary insomnia through 60 sleep-related items. To shorten this questionnaire without sacrificing accuracy, Lee et al. [43] applied a random forest algorithm to clinical data (*n* = 310), resulting in a shortened version known as MCQI-6. In this process, feature importance was calculated using the mean decrease in accuracy method, which measures the reduction in the model’s accuracy when each question is removed. To ensure stable rankings, the results from 500 different random forest models were averaged. Based on these feature importance scores, the top six key questions were selected to distinguish between individuals with normal sleep patterns and those suffering from insomnia. This is achieved by performing a regression task to estimate the total sum of responses from the original 60 questions. MCQI-6 has demonstrated a good fit, with a Cronbach’s *α* of 0.96, and has shown good internal consistency (*α* = 0.843).

The study protocol was approved by the Institutional Review Board of Sungshin Women’s University, Seoul, South Korea (SSWUIRB-2020-009). Written informed consent was waived. The survey was administered anonymously, and no personal information was gathered. This survey form was developed according to the Checklist for Reporting Results of Internet e-Surveys (CHERRIES) guidelines [57].

#### 1.1.2. Machine learning-based shortened questionnaire, SLEEPS

SLEEPS is a straightforward questionnaire designed to predict sleep disorders using XGBoost models [25]. From an initial set of 30 features consisting of 22 items from sleep-related questions and 8 demographic characteristics, feature importances were calculated using the absolute SHAP values, which measure each feature’s contribution to the model’s predictions. To ensure stability, the results from multiple iterations of the XGBoost model were averaged. Based on these SHAP importance scores, 9 key questions were selected to calculate the risk of three sleep disorders—insomnia, comorbid insomnia and sleep apnea (COMISA), and obstructive sleep apnea (OSA)—without a complex polysomnography (PSG) test. For all three sleep disorders, SLEEPS shows high accuracy (AUROC 0.9). Furthermore, a publicly accessible website (https://sleep-math.com) based on this algorithm has been created, allowing individuals to easily predict their risk of these conditions.

The study protocol was reviewed and approved by the Institutional Review Board of the SMC (approval 2022-07-003) and was conducted in accordance with the principles of the Declaration of Helsinki. Participant informed consent was waived due to the retrospective nature of the study.

#### 1.1.3. Current approaches for shortened questionnaires

In various healthcare settings, questionnaires with *m* questions 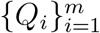 are utilized to predict the risk severity of a disease based on the responses 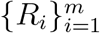 (Fig. 1 (a)). However, extensive questionnaires can lead to lower response rates and incomplete submissions. To mitigate these issues, shortened versions of questionnaires 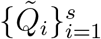, where *s < m*, subset of 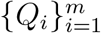, have been developed using machine learning techniques [25, 30, 40, 41, 42, 43, 44, 45, 46, 47, 48, 49, 58].

**Figure 1.**
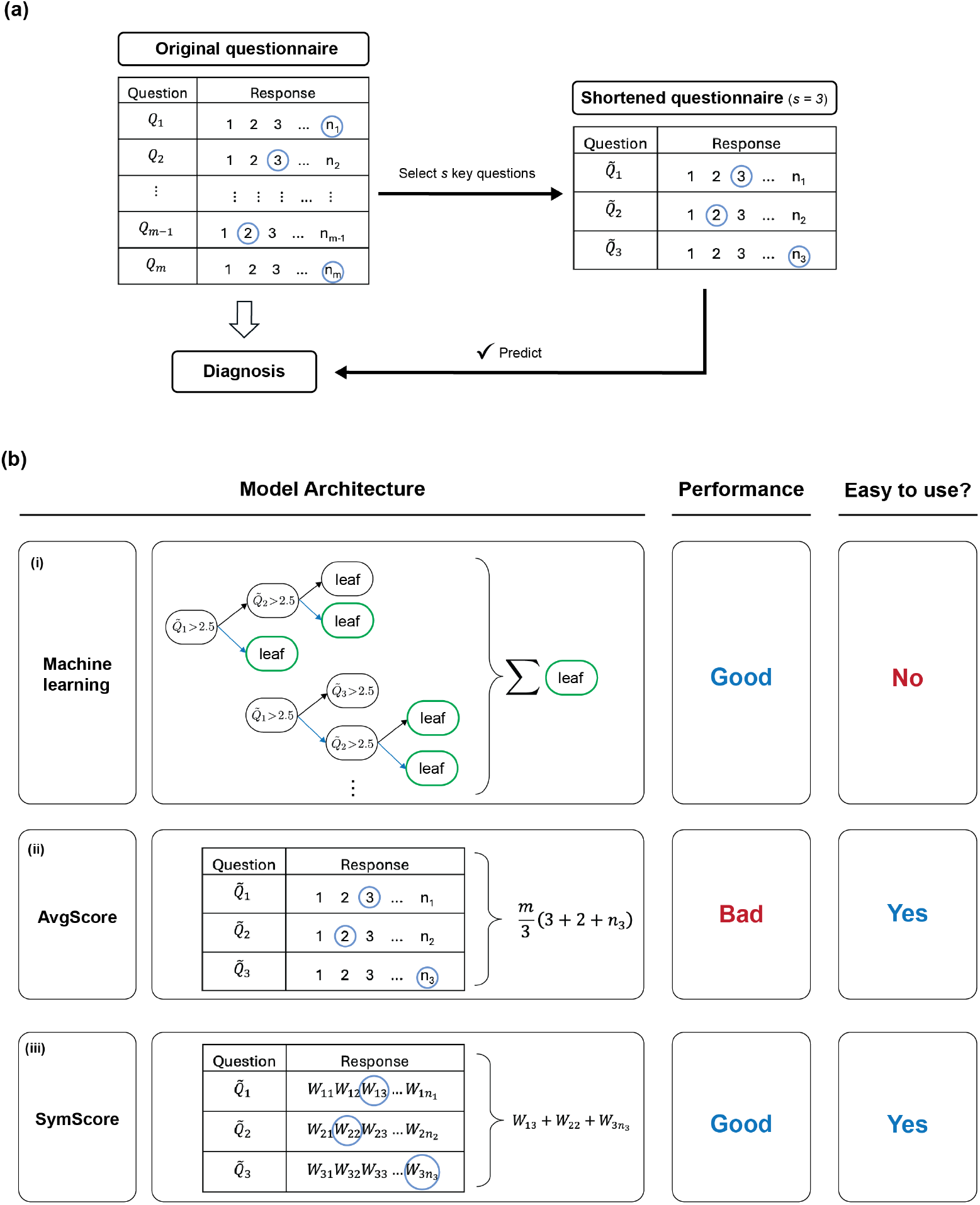
Problem formulation and approaches. (a) Original questionnaires consist of *m* questions with *n*_*i*_ (*i* = 1,…, *m*) possible responses for each question. A subset of *s* key questions with *s < m* is selected to shorten the questionnaire while maintaining diagnostic accuracy similar to that of the original questionnaire. (b) For this task, three approaches are employed: (i) Machine learning-based approaches, (ii) AvgScore, and (iii) SymScore. While the machine learning-based approach is accurate, it is limited by its black-box nature, making it difficult to use. AvgScore simplifies the process by simply scaling the average response of the shortened questionnaire by 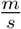, but it tends to yield lower accuracy. SymScore ensures accuracy by assigning different weights (*W*_*ij*_) to each response, while also maintaining ease of use.

Subsequently, machine learning algorithms are used to predict disease severity by estimating the sum of the responses 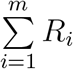 *R*_*i*_ from the original questionnaires. They utilize the responses 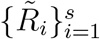 to the shortened questionnaires to achieve these predictions as follows:

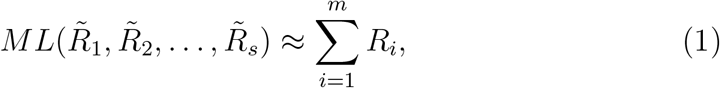

where *ML* represents a machine learning model. However, due to its blackbox nature, the mathematical expression of the machine learning model described in Eq. 1 is not explicitly expressed, making it difficult to interpret the impact of each response on the disease severity. Furthermore, the required computational demands and specialized knowledge limit their practical use in clinical settings (Fig. 1 (b) (i)).

Because machine learning-based shortened questionnaires are difficult to use in clinical settings, the sum of the responses from a shortened questionnaire scaled by the ratio 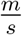 is often heuristically utilized in real clinical settings as follows:

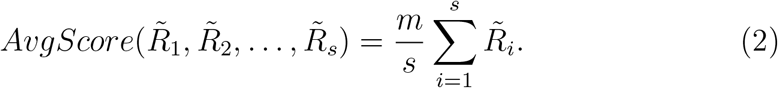

This simple rescaling method, while easier to implement, is less likely to maintain the prediction accuracy of the original machine learning-based approach (Fig. 1 (b) (ii)).

## 2. Methods

To address both the accuracy limitations of heuristic averaging approaches and the lack of interpretability of machine learning models, we developed SymScore. SymScore uses the following weighted sum of responses to predict disease severity:

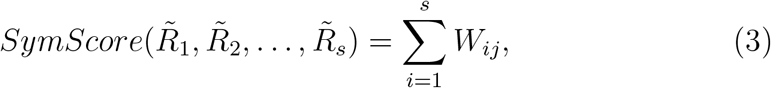

if 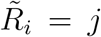 is the response to question 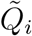 (Fig. 1 (b) (iii)). Thus, all responses have their own weights *W*_*ij*_ to predict the severity of the disease, yielding more accurate predictions than AvgScore. By using these weights, we can directly generate a score table where each response is associated with a specific weight (Fig. 1 (b) (iii) left). This method allows us to obtain the interpretable shortened questionnaires, unlike machine learning-based models that are inexplicable. Furthermore, the sum of the weights *W*_*ij*_ leads to the prediction of disease severity, which is much simpler than machine learning approaches (Fig. 2 (a)).

**Figure 2.**
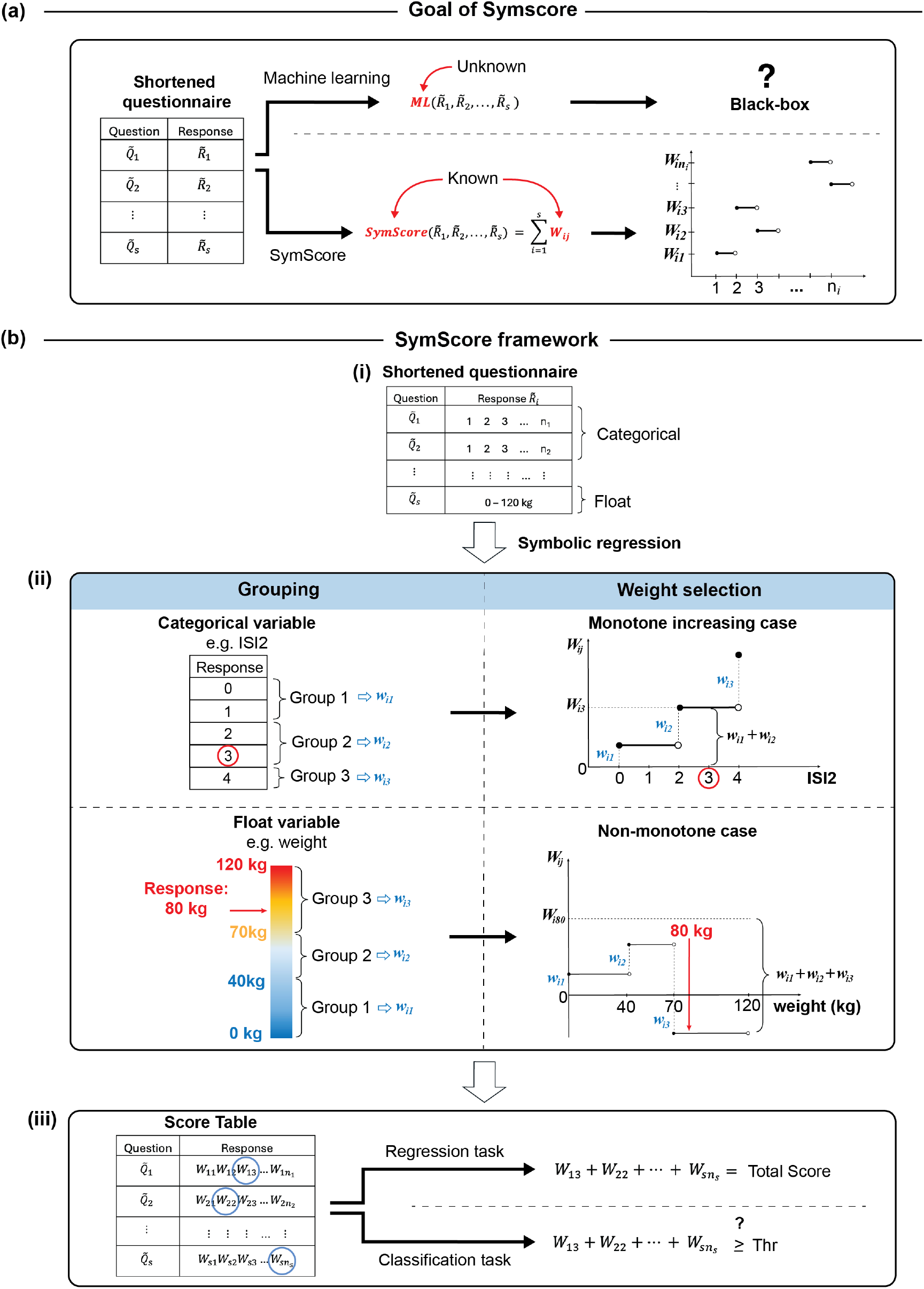
Goal and framework of SymScore for producing interpretable shortened questionnaires. (a) From the shortened questionnaire with *s* questions, machine learning-based approaches represent disease severity as a function of the responses to each question (ML). This function is frequently complex and not explicitly defined. In contrast, SymScore represents disease severity as the sum of weights for each response, making the model interpretable and computationally efficient. (b) (i) The shortened questionnaire includes questions that require responses in either categorical or float values. (ii) SymScore automatically groups these responses and assigns weights based on their importance. For categorical variables (e.g., ISI2), where severity typically increases with the response, the weights *W*_*ij*_ assigned for each response should increase. For this, the partial weights *w*_*ik*_, whose sum determines *W*_*ij*_, are restricted to be nonnegative. For float variables, such as weight, the weights *W*_*ij*_ may not be monotonic, allowing for negative partial weights. (iii) Based on the weights (*W*_*ij*_) assigned to each response, a score table is produced. The total score is computed by simply summing the weights of responses. For regression tasks, the total score assesses disease severity, while for classification tasks, the total score is compared with a threshold to determine whether an individual has a certain disease.

We designed SymScore (Fig. 2 (b) and Algorithm 1) to automatically assign the weights *W*_*ij*_ to the responses of the shortened questionnaire. SymScore consists of the following modules.

### 2.1. Module 1: Grouping question responses

The first module focuses on grouping question responses to the shortened questionnaire. The shortened questionnaire may consist of questions that obtain either categorical or float values as responses (Fig. 2 (b) (i)). For the computational efficiency of SymScore, we categorize these responses into groups having the same weight (Fig. 2 (b) (ii) left and Table 1). We denote these groups by *G*_*ik*_, with *i* = 1,…, *s*, and *k* = 1,… *g*_*i*_, where *g*_*i*_ is the number of response groups for question *Q*_*i*_. To perform the grouping during the fitting, the loss function becomes discontinuous as the grouping changes, and thus typical regression methods cannot be used. To address this, we utilize symbolic regression, which does not require continuity in the loss function. Symbolic regression is particularly well-suited for this task because of its ability to determine the optimal number of response groups. While increasing the number of response groups can improve prediction precision, it also increases the risk of overfitting and can reduce computational efficiency. Symbolic regression balances this trade-off by identifying the minimum number of response groups necessary without compromising accuracy. Details on the optimal response grouping process can be located in the Supplementary Material (S3) and is illustrated in Fig. S1.

**Table 1:** Illustration of partial and total weights for each response in the shortened questionnaire produced by SymScore. The table demonstrates the process of grouping responses and assigning weights within the SymScore framework. For each question 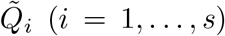, the responses 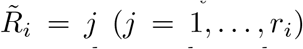 are grouped into *G*_*ik*_ (*k* = 1,…, *g*_*i*_), with each group having an assigned partial weight *w*_*ik*_. The summation of the partial weights *w*_*i*1_, *w*_*i*2_,…, *w*_*ik*_ gives the total weight *W*_*ij*_ assigned to each response *j*. This systematic grouping and assigning of weights forms the score table, enabling the summation of responses to facilitate disease diagnosis.

| Question | Response | Group | Partial weight | Total weight |
| --- | --- | --- | --- | --- |
| $\tilde{Q}_1$ | 1 | $G_{11}$ | $w_{11}$ | $W_{11} = w_{11}$ |
| | 2 | | | $W_{12} = w_{11}$ |
| | 3 | $G_{12}$ | $w_{12}$ | $W_{13} = w_{11} + w_{12}$ |
| | 4 | | | $W_{14} = w_{11} + w_{12}$ |
| | 5 | $G_{13}$ | $w_{13}$ | $W_{15} = w_{11} + w_{12} + w_{13}$ |
| $\tilde{Q}_2$ | 1 | $G_{21}$ | $w_{21}$ | $W_{21} = w_{21}$ |
| | 2 | | | $W_{22} = w_{21}$ |
| | 3 | | | $W_{23} = w_{21}$ |
| | 4 | $G_{22}$ | $w_{22}$ | $W_{24} = w_{21} + w_{22}$ |
| ⋮ | ⋮ | ⋮ | ⋮ | ⋮ |
| $\tilde{Q}_s$ | 0 | $G_{s1}$ | $w_{s1}$ | $W_{s1} = w_{s1}$ |
| | 1 | | | $W_{s2} = w_{s1}$ |
| | 2 | $G_{s2}$ | $w_{s2}$ | $W_{s3} = w_{s1} + w_{s2}$ |
| | 3 | $G_{s3}$ | $w_{s3}$ | $W_{s4} = w_{s1} + w_{s2} + w_{s3}$ |
| | 4 | | | $W_{s5} = w_{s1} + w_{s2} + w_{s3}$ |

### 2.2. Module 2: Partial weights technique

In addition to identifying the optimal number of response groups, SymScore also estimates the weights *W*_*ij*_ given in Eq. 3 for each of the *i*-th responses 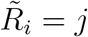 in the shortened questionnaire, such that their sum closely approximates the sum of the responses *R*_*l*_ in the original questionnaire. Specifically,

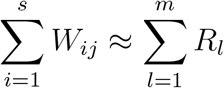

To determine the weights for the response 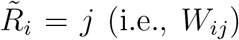, we divide each *W*_*ij*_ into partial weights 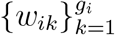, where each partial weight is associated with a response group *G*_*ik*_. These partial weights are added to compute the total weight *W*_*ij*_ (Table 1). Specifically, if the response *j* for the *i*-th question is in the *k*-th group *G*_*ik*_, then the total weight is given by

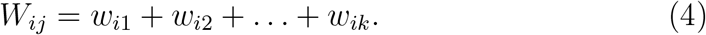

For instance, in Fig. 2 (b) (ii), since the chosen response is *j* = 3, which is in *G*_*i*2_, the weight *W*_*i*3_ is computed as *W*_*i*3_ = *w*_*i*1_ + *w*_*i*2_. Similarly, if the chosen response is *j* = 2, which is also in *G*_*i*2_, then *W*_*i*2_ = *w*_*i*1_ + *w*_*i*2_. Therefore, the weight *W*_*ij*_ is the same for responses that are in the same group. This grouping of responses with the same weight enhances fitting efficiency.

The weights *W*_*ij*_ often increase as *j* increases when the response value increases with severity level. For the monotone increasing responses, the partial weights are restricted to be nonnegative, ensuring that *W*_*ij*_ increases as *j* increases (Fig. 2 (b) (ii) upper right). On the other hand, for non-increasing responses, negative partial weights are allowed (Fig. 2 (b) (ii) lower right).

### 2.3. Module 3: Training SymScore

We obtain the optimal partial weights *w*_*ik*_ for each response group through the following steps:

1. 20,000 populations of *{w*_*ik*_*}* are randomly generated using symbolic regression, forming the first generation.
2. The performance of each population in the generation is evaluated by comparing the true target values with the predicted values, using the mean absolute error (MAE) as the fitness metric.
3. The top 30% of the population with the lowest MAE are selected, and from these selected populations, a new generation is then created. For every new generation created, the grouping of responses is changed during the fitting process.

This process of generating new populations and evaluating their performance is repeated until no further improvement in predictive performance is observed. This iterative process leads to the identification of the optimal grouping and partial weights *w*_*ik*_, characterized by the highest predictive performance. Details of the parameter tuning process for training the symbolic regression model can be found in the Supplementary Material (S1).

### 2.4. Module 4: Predicting disease severity using the computed weights

Using the partial weights *w*_*ik*_, the total weight *W*_*ij*_ for each response is calculated (Fig. 2 (b) (ii) right). By summing all *W*_*ij*_, SymScore accurately predicts the total score of the original questionnaire (Fig. 2 (b) (iii)). This total score estimates the disease severity in both regression and classification tasks.

#### Regression tasks

The severity of the disease was originally predicted using the total score of the original questionnaire. Thus, for regression tasks, the maximum total value of the shortened questionnaire must be the same as the maximum total value of the responses from the original questionnaire. In other words, the weights *W*_*ij*_ need to satisfy the following condition:

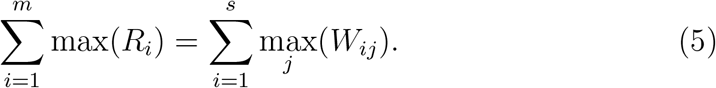

To meet the above constraint, we generated artificial data points where each response *R*_*i*_ is set to its maximum value and the corresponding target value is set to the maximum total value from the original questionnaire, i.e., 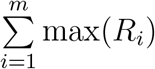. These artificial data are appended to the training set, creating a new augmented dataset. This dataset is subsequently used for training, following the process described in Module 3. Incorporating artificial data into the training set helps the model learn to predict the maximum total score effectively.

However, it is important to note that this method of adding artificial data does not always guarantee that the constraint given in Eq. 5 is satisfied. If the difference 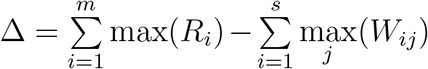 is positive (i.e., Δ *>* 0), one of the weights *w*_*ik*_ is increased by 6. This weight is then used to estimate performance by comparing the sum of the original responses and the predicted values. This procedure is repeated for all possible weights, and the weight with the highest prediction performance is selected. These adjusted partial weights are then summed to compute the total weight *W*_*ij*_, which is used to construct the score table. Summing the total weights *W*_*ij*_ (*i* = 1,…, *s*) provides the total score, which is used to predict disease severity for regression tasks.

#### Classification tasks

For classification tasks, a threshold for the sum of the weights *W*_*ij*_ is necessary to distinguish between individuals with and without the disease. This threshold is determined by defining a probability *p* that indicates the likelihood of an individual having the disease. The probability *p* is calculated using the sum of the weights *W*_*ij*_ and a constant bias *b* as follows:

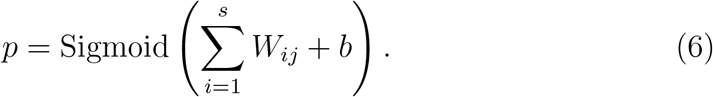

Using this probability, SymScore is trained according to the method outlined in Module 3. As a result, we obtain the weights *W*_*ij*_ and the constant *b*. Subsequently, we obtain a classification threshold for 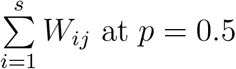 as follows:

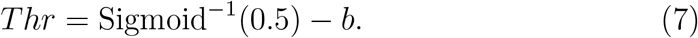

Based on this threshold, we can classify the participants into two groups: those who satisfy 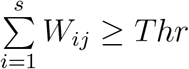 and those who do not. For example, in questionnaires used to assess a certain disease, patients with a total score above the threshold are classified as having the disease. Thus, this comparison to the threshold is the final step in using the SymScore-derived questionnaire for disease assessment. More technical details for the threshold computation can be found in the Supplementary Material (S4).

### 2.5. Model Evaluation and Data Handling

To evaluate the predictive performance of the shortened questionnaire derived from SymScore, we compared it with other existing methods. To achieve this, we randomly selected 30% of the entire dataset as the test set for measuring the performance of the shortened questionnaire. This process was repeated for 10,000 different shufflings of the test set for robustness. Subsequently, we calculated performance metrics for each shortened questionnaire on every test set. For regression tasks, we utilized MAE and coefficient of determination *R*^2^. For classification tasks, we used the area under the receiver operating characteristic curve (AUROC) as a performance metric.

Since data imbalance can affect the performance of the model, we addressed this issue in the SymScore code by creating a balanced data set. Specifically, a dataframe was created consisting of all rows where patients were diagnosed with the sleep disorder, along with another dataframe containing an equal number of rows where patients did not have the disorder. This ensures that the number of negative samples is precisely balanced with the number of positive ones. Then, the SymScore algorithm was applied to this dataset.

Moreover, when splitting the data into training and test sets, the `stratify` argument was employed to maintain class balance in both subsets. This guarantees that both the training and testing datasets contain an equal proportion of positive and negative cases. By first creating a balanced dataset and then stratifying the train-test split, this approach effectively addresses the issue of imbalanced data, reducing the likelihood of the model being biased toward the majority class and enhancing the overall fairness and accuracy of the predictions.

To address potential overfitting issues in regression tasks, two strategies were implemented to ensure the model’s robustness and generalization. The model’s performance on both the training and the testing sets was evaluated using the Mean Absolute Error (MAE). A threshold ratio was then established to compare the test MAE against the training MAE. If the test MAE exceeds 1.5 times the training MAE, a warning is issued indicating potential overfitting. Potential overfitting in the classification tasks is addressed in the same manner, but using AUROC as the performance metric.

To further assess the model’s performance in regression tasks, K-fold cross-validation with five splits was employed. This technique enhances model evaluation by dividing the data into training and validation subsets, ensuring that each data point is used for both training and validation across different iterations. The mean and standard deviation of the cross-validated MAE were computed to assess the generalization capability of the model. if the mean cross-validated MAE is more than 1.5 times the training MAE, a warning of potential overfitting is issued. Additionally, if the cross-validated MAE’s standard deviation exceeds 15% of the mean cross-validated MAE a, warning of the model’s instability is provided. The assessment of the model’s performance in classification tasks is similarly measured, but with AUROC as the performance metric.

Through these comprehensive steps, the analysis aimed to ensure that the model generalizes well to unseen data and remains robust against overfitting, thereby providing a reliable prediction framework for the study.

#### Algorithm 1

SymScore (Symbolic Regression-based Clinical Score Generator)

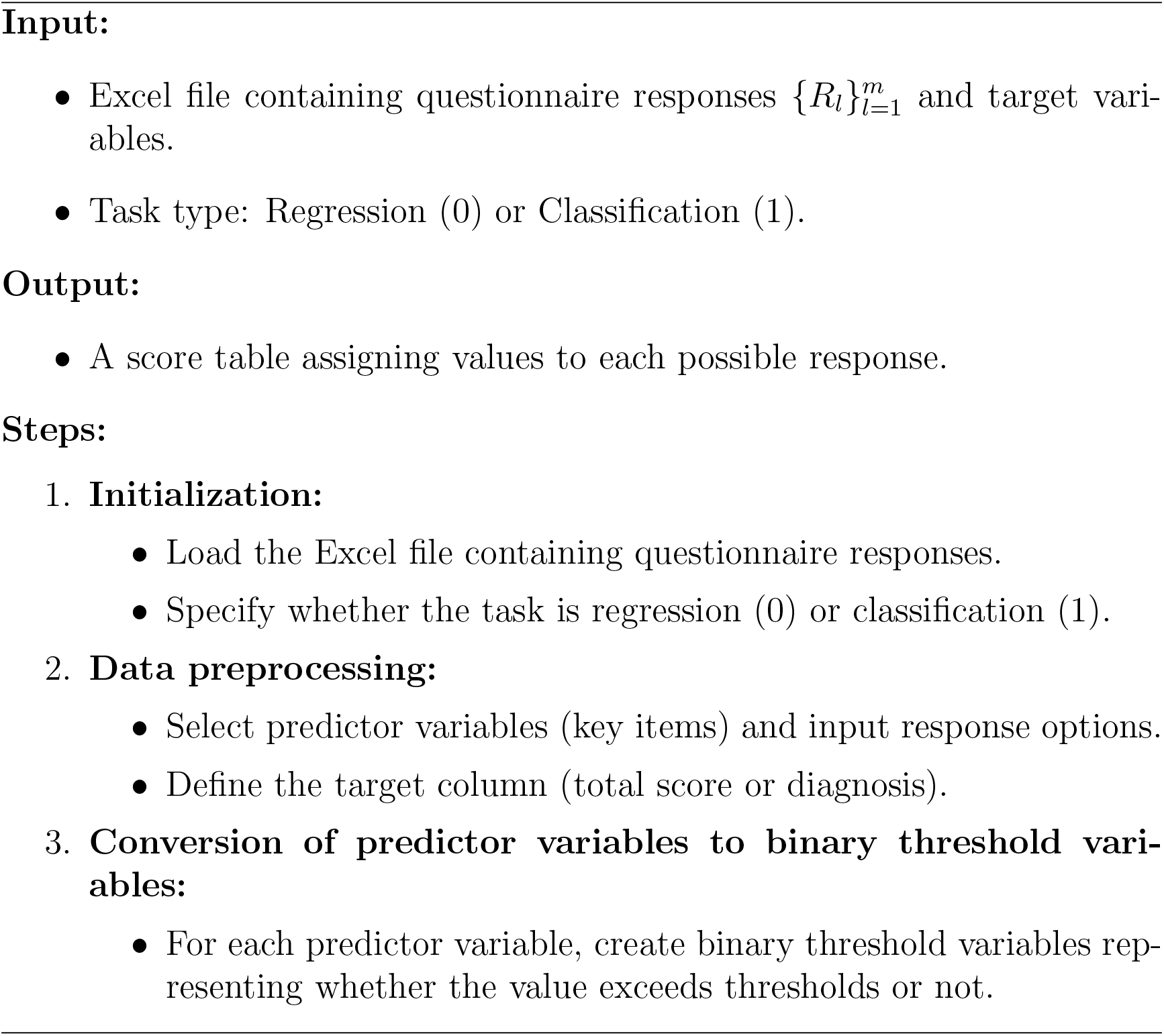
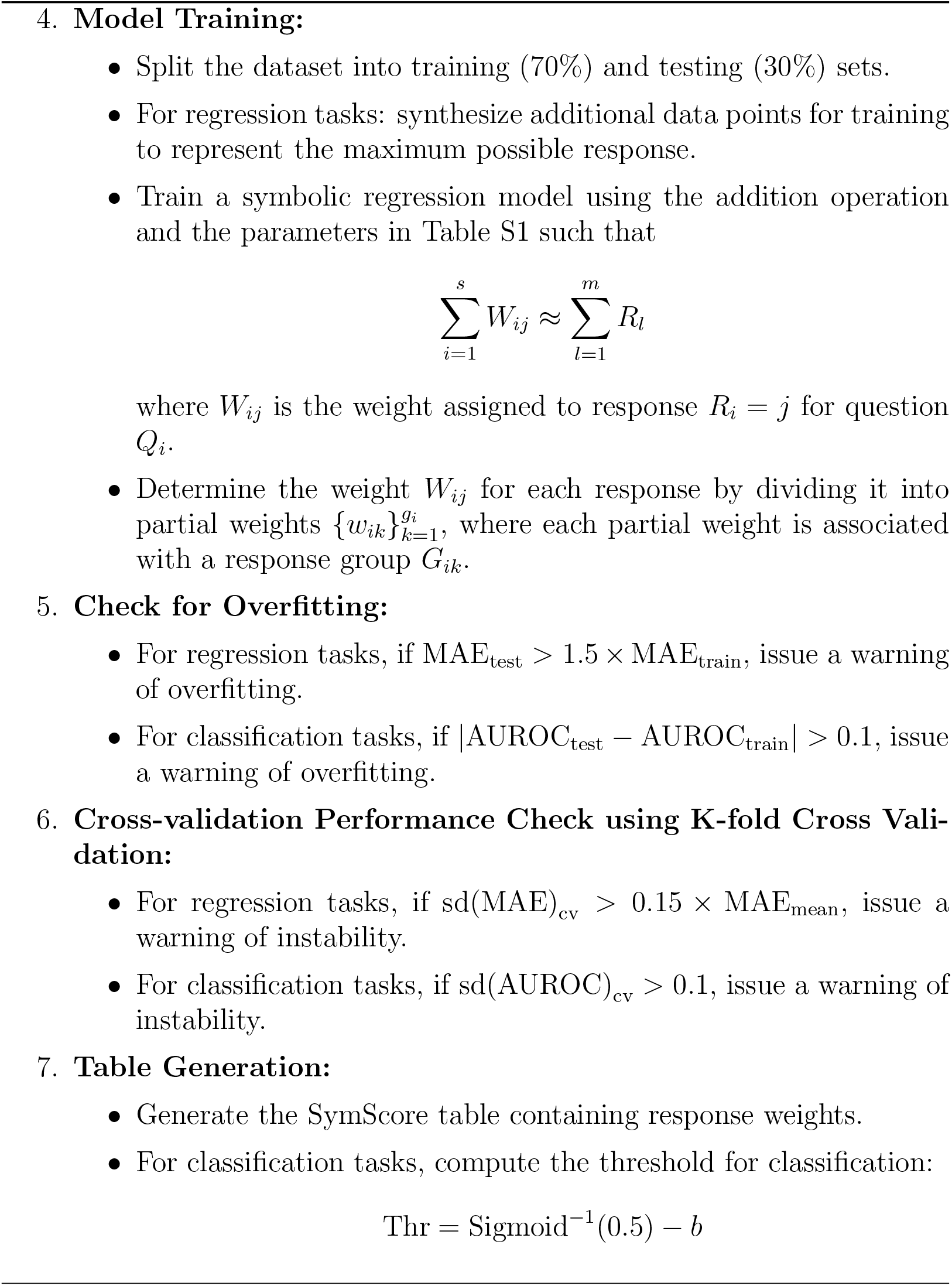

## 3. Results

### 3.1. SymScore can generate an easy-to-use shortened questionnaire of MCQ-I for assessing metacognitive beliefs related to insomnia

The Metacognitions Questionnaire-Insomnia (MCQ-I) was created to assess metacognitive beliefs in individuals with primary insomnia [59]. Each individual rates 60 questions on a four-point Likert scale, and the total sum is used to evaluate the extent and impact of maladaptive metacognitive beliefs related to insomnia.

To reduce the time needed for this assessment, Lee et al. [43] used a random forest algorithm on clinical data to identify six key questions (MCQI-6) based on feature importance. Using MCQI-6, they performed a regression task to estimate the total sum of responses from the MCQ-I questions. Although this method effectively reduced the number of questions without sacrificing accuracy, its practical use is limited by the need for computational resources and machine learning expertise among healthcare staff. As a workaround, the scaled average value of responses from the six key questions was heuristically used, which we referred to as AvgScore (Fig. 1 (b) (ii)).

To address this issue, we used SymScore to create a simple scoring table that allows easy summation without computers while maintaining predictive performance (Table 2). We evaluated this SymScore shortened questionnaire along with the MCQI-6 and AvgScore for predicting the MCQI-60 total score using the six key questions. For this evaluation, we randomly selected 30% of the clinical dataset as the test set, and used a scatter plot to visualize the prediction results (Fig. 3 (a) and Table 3). The scatter plots for MCQI-6 and SymScore show points tightly clustered along the diagonal line, indicating good predictive performance. In contrast, the AvgScore scatter plot exhibits a broader dispersion of points around the diagonal, suggesting lower predictive accuracy. Additionally, AvgScore tends to underestimate low measured values and overestimate high measured values. This is the result of simply scaling the average of the responses of the shortened questionnaire.

**Table 2:**
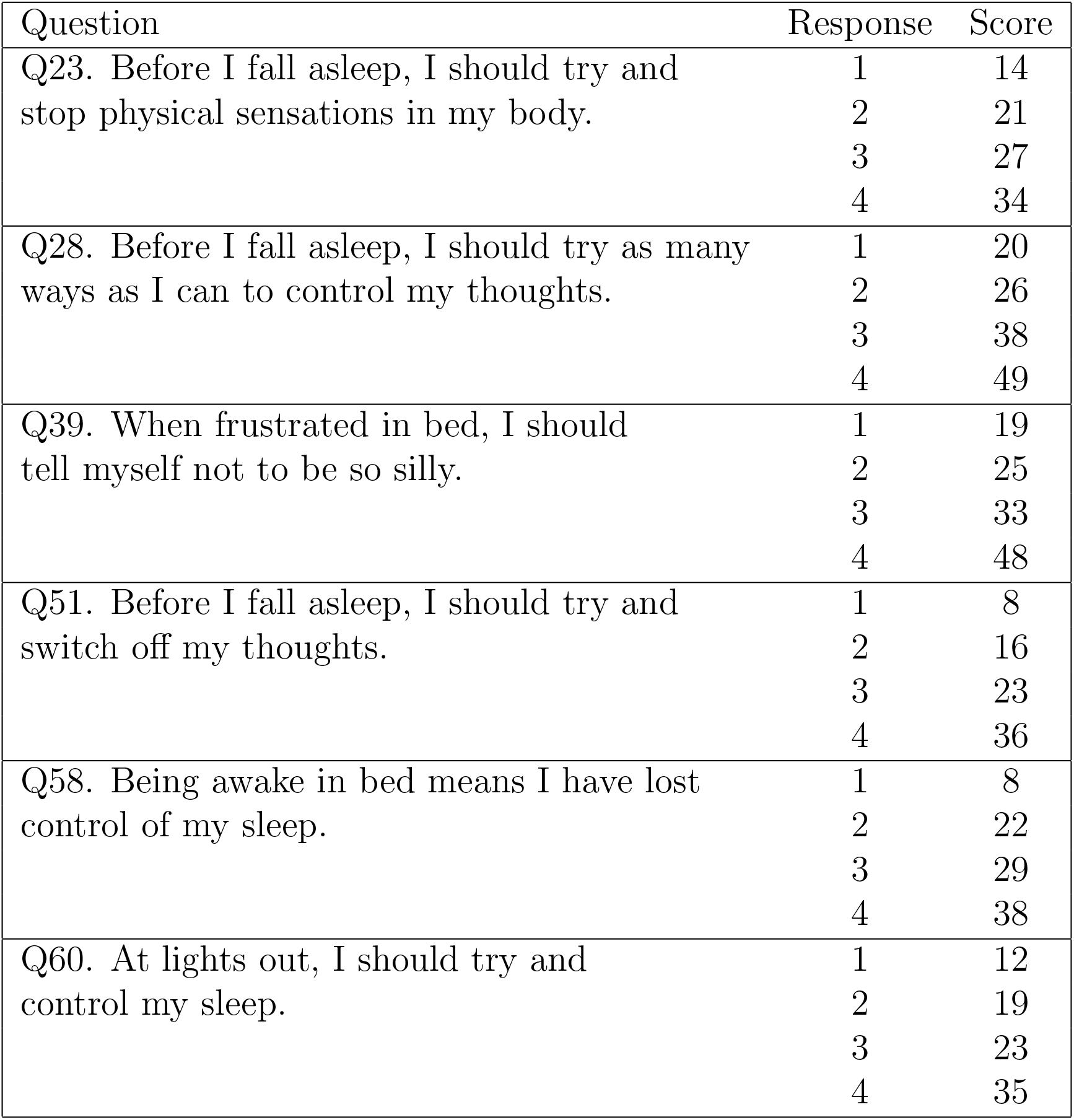
Simplified score table of MCQI-6 using SymScore. Responses range from 1 (Do not agree) to 4 (Agree very much), corresponding to increasing degrees of agreement. Summing the score of responses for all questions helps assess metacognitive beliefs related to insomnia.

| Question | Response | Score |
| --- | --- | --- |
| Q23. Before I fall asleep, I should try and stop physical sensations in my body. | 1 | 14 |
|  | 2 | 21 |
|  | 3 | 27 |
|  | 4 | 34 |
| Q28. Before I fall asleep, I should try as many ways as I can to control my thoughts. | 1 | 20 |
|  | 2 | 26 |
|  | 3 | 38 |
|  | 4 | 49 |
| Q39. When frustrated in bed, I should tell myself not to be so silly. | 1 | 19 |
|  | 2 | 25 |
|  | 3 | 33 |
|  | 4 | 48 |
| Q51. Before I fall asleep, I should try and switch off my thoughts. | 1 | 8 |
|  | 2 | 16 |
|  | 3 | 23 |
|  | 4 | 36 |
| Q58. Being awake in bed means I have lost control of my sleep. | 1 | 8 |
|  | 2 | 22 |
|  | 3 | 29 |
|  | 4 | 38 |
| Q60. At lights out, I should try and control my sleep. | 1 | 12 |
|  | 2 | 19 |
|  | 3 | 23 |
|  | 4 | 35 |

**Table 3:** Comparison of performance among MCQI-6 (ML-based), AvgScore, and SymScore.

|  | MCQI-6 | AvgScore | SymScore |
| --- | --- | --- | --- |
| MAE | 9.94 | 14.74 | 10.73 |
| $R^2$ score | 0.82 | 0.58 | 0.77 |

**Figure 3.**
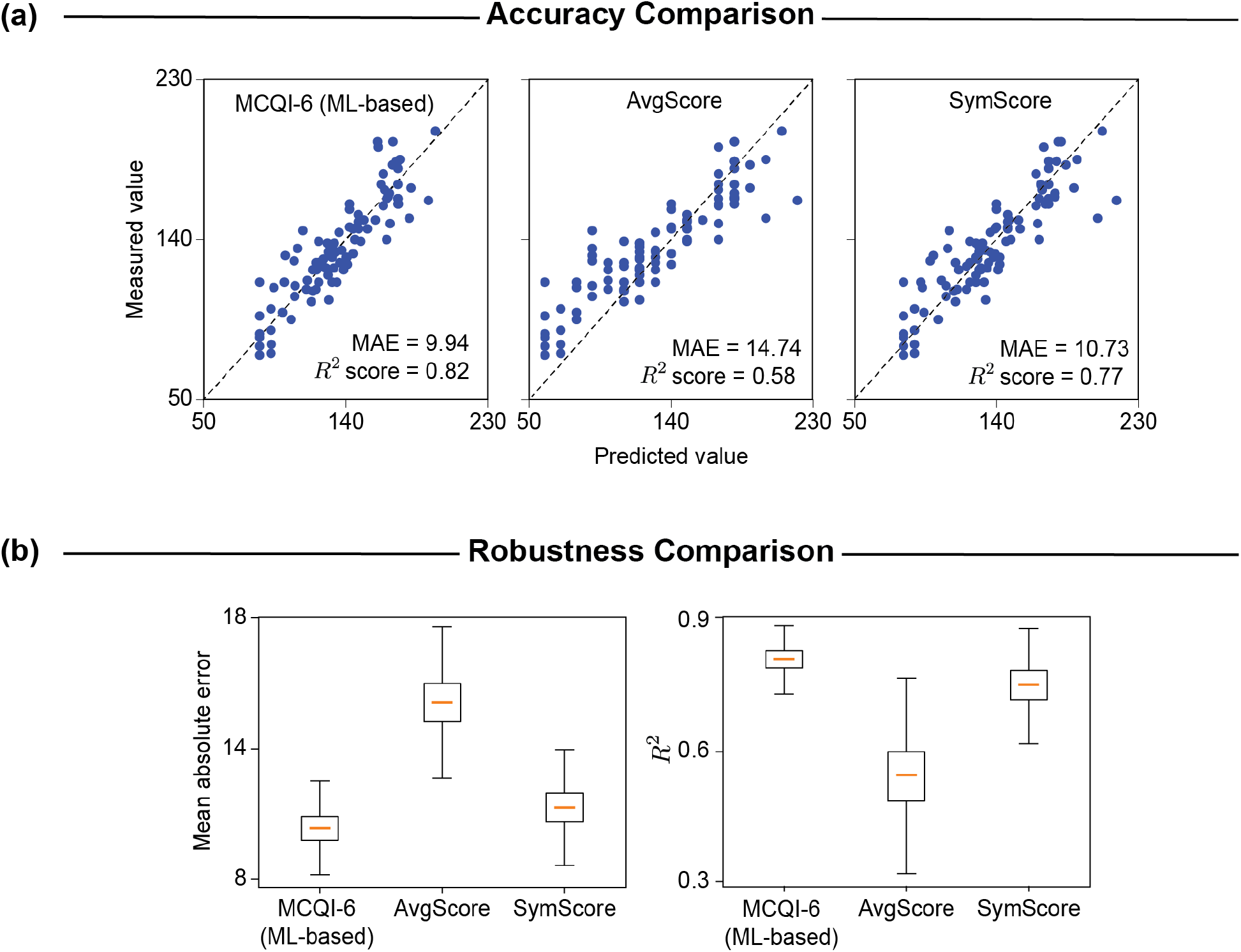
Evaluating the predictive performance of three shortened questionnaires based on MCQI-6 using machine learning, AvgScore, and SymScore. (a) The scatter plots of predicted scores versus measured scores for the 93 participants in the test set show that MCQI-6 and SymScore (Table 2) are more closely clustered around the diagonal line compared to AvgScore, indicating better agreement between predicted and measured scores for these two methods. (b) Compared to MCQI-6 using machine learning, shortened questionnaires generated with SymScore (Table 2), but not AvgScore, yield comparable performances.

Furthermore, we employed two metrics for quantitative assessment: MAE and coefficient of determination (*R*^2^) (Fig. 3 (b)). We verified that the model was not overfitting by comparing its performance on both the training and the testing sets (Table S4). Additionally, we assessed the model’s stability and generalizability through K-fold cross-validation across different data subsets. To ensure robustness, we repeated the evaluation process 10,000 times with different test set shuffling. Our evaluation showed that compared to the MCQI-6 based on a random forest algorithm (MAE = 9.94, *R*^2^ = 0.82), the AvgScore shortened questionnaire exhibited lower performance (MAE = 14.74, *R*^2^ = 0.58). On the other hand, the SymScore shortened questionnaire (MAE = 10.73, *R*^2^ = 0.77) achieved performances comparable to the MCQI-6. Overall, these results highlight that MCQI-6 and SymScore have comparable predictive performances, both being superior to AvgScore.

Importantly, SymScore provides a simple and transparent score table (Table 2). This table can be used as a basic summative questionnaire, allowing for the direct addition of weights without the necessity of computers, in contrast to the MCQI-6. The transparency of the SymScore table enables easy interpretation and analysis of prediction results. The scoring table reflects the monotonicity of weights, capturing an increasing order of risk severity. This design ensures that higher severity responses receive higher weights, facilitating a nuanced understanding of risk associated with each response. The differentiation in weight scales across questions quantifies each question’s contribution to the total score prediction. For instance, questions Q28 and Q39 have the highest weights (48-49), indicating their greater importance compared to others with weights between 34 and 38. This variance highlights the questionnaire’s ability to differentiate the severity or risk associated with each of the responses. Thus, SymScore offers a simple and interpretable shortened questionnaire that can be easily implemented in medical centers.

### 3.2. SymScore can produce short and interpretable questionnaires for diagnosis of sleep disorders

SLEEPS is an XGBoost-based algorithm designed to predict the risk of OSA, insomnia, and COMISA using responses to nine straightforward questions, eliminating the need for complex PSG tests. While SLEEPS demonstrates high predictive performance with AUROC scores of 0.88 for OSA, 0.93 for insomnia, and 0.94 for COMISA on a test set comprising 30% of the data (Fig. 4 (a)-(c)), its clinical adoption has been limited due to the inherent lack of interpretability in machine learning models.

**Figure 4.**
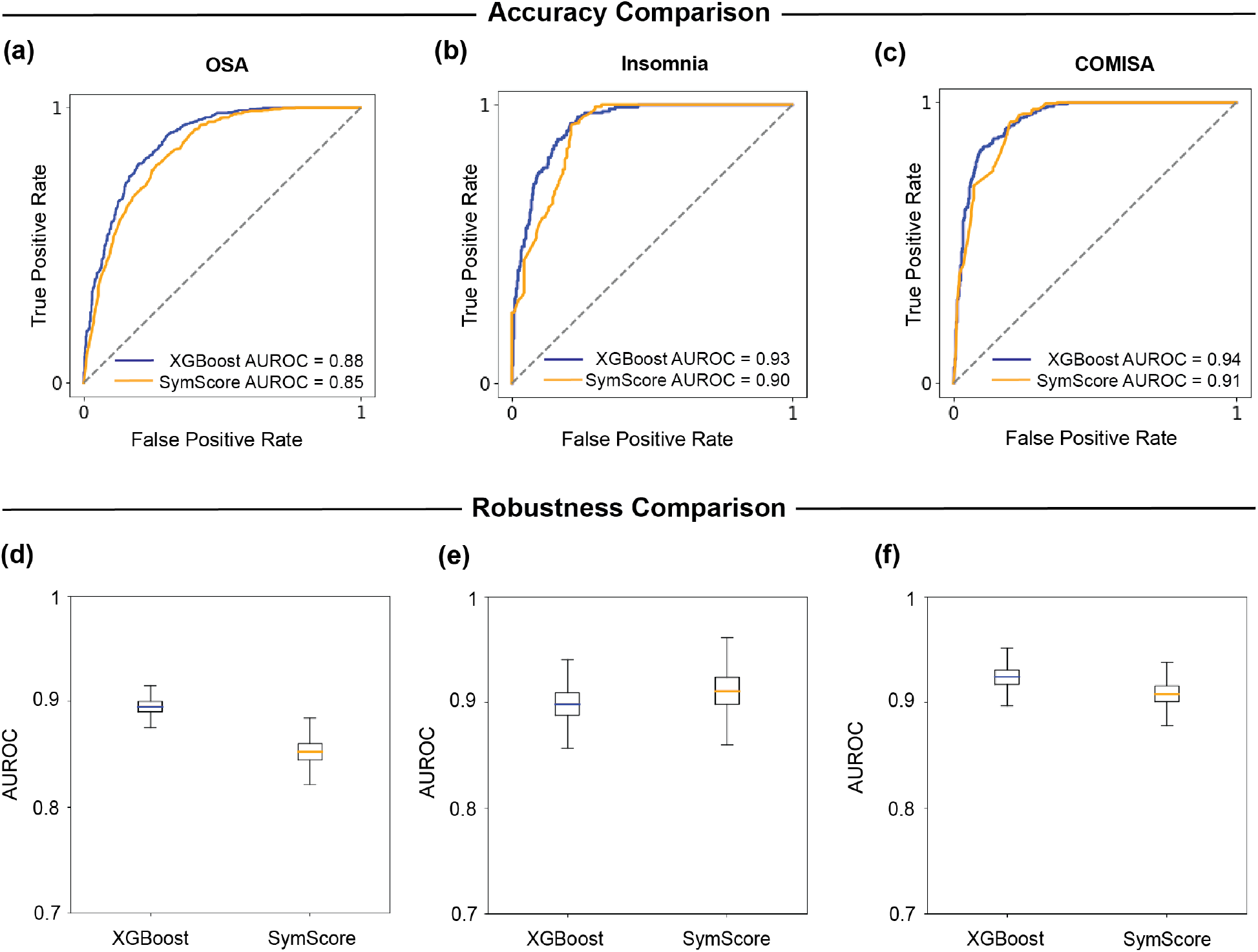
Comparison of performance between SLEEPS based on XGBoost and the shortened questionnaire based on SymScore for insomnia, OSA, and COMISA. (a)-(c) Area under the receiver operating curve (AUROC) of predictions for the test set from XGBoost and SymScore for OSA (Table 6 column 2), insomnia (Table 6 column 3), and COMISA (Table 6 column 4) are comparable in performance. (d)-(f) For 10,000 different test set shufflings, XGBoost and SymScore (Table 6) demonstrate comparable predictive performance for diagnosing OSA, insomnia, and COMISA.

To address this challenge, we developed a simplified questionnaire using SymScore (Table 6) and evaluated its performance using AUROC. The SymScore questionnaires displayed satisfactory performance, with AUROC scores of 0.85 for OSA, 0.90 for insomnia, and 0.91 for COMISA (Fig. 4 (a)-(c) and Table 4). Moreover, SymScore’s performance remained comparable to that of XGBoost even after repeating the evaluation process 10,000 times with different test set shuffling (Fig. 4 (d)-(f)).

**Table 4:** Comparison of performance between XGBoost and SymScore.

|  |  | XGBoost | SymScore |
| --- | --- | --- | --- |
| AUROC | OSA | 0.88 | 0.85 |
|  | Insomnia | 0.93 | 0.90 |
|  | COMISA | 0.94 | 0.91 |

We compare SymScore’s performance only with XGBoost, not AvgScore, due to the nature of the SLEEPS questionnaire. Unlike other questionnaires where AvgScore can predict total scores by rescaling, this isn’t feasible here because of float variables like weight and BMI. Specifically, in the SLEEPS questionnaire, categorical variables (0-4 scale) and float variables (30-181 kg) differ in response scales, making a uniform scaling factor inapplicable. Therefore, we compare SymScore’s performance only with XGBoost.

To enhance clinical relevance, we include sensitivity and specificity values, which are often more informative in clinical contexts than metrics like MAE and AUROC (Table 5). We determined the optimal thresholds by minimizing the false positive rate (FPR) and maximizing the true positive rate (TPR), yielding the sensitivity and specificity values for each sleep disorder. Comparing SymScore with XGBoost, SymScore demonstrates comparable, if not superior, performance in correctly identifying individuals with sleep disorders. Specifically, SymScore achieves higher sensitivity for OSA (0.92 vs. 0.90) and COMISA (0.93 vs. 0.92), while maintaining a sensitivity of 0.90 for insomnia, below the XGBoost sensitivity value of 0.92.

**Table 5:**
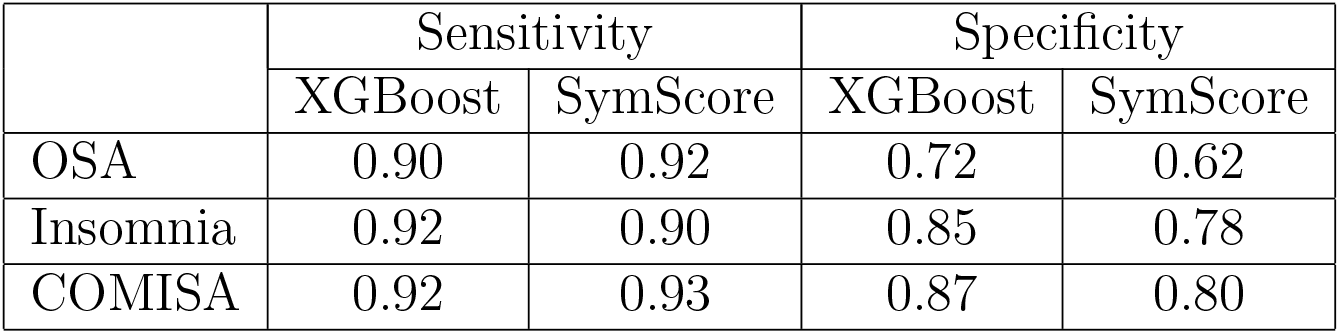
Comparison of sensitivity and specificity of the models derived from XGBoost and SymScore.

|  | Sensitivity |  | Specificity |  |
| --- | --- | --- | --- | --- |
|  | XGBoost | SymScore | XGBoost | SymScore |
| OSA | 0.90 | 0.92 | 0.72 | 0.62 |
| Insomnia | 0.92 | 0.90 | 0.85 | 0.78 |
| COMISA | 0.92 | 0.93 | 0.87 | 0.80 |

**Table 6:**
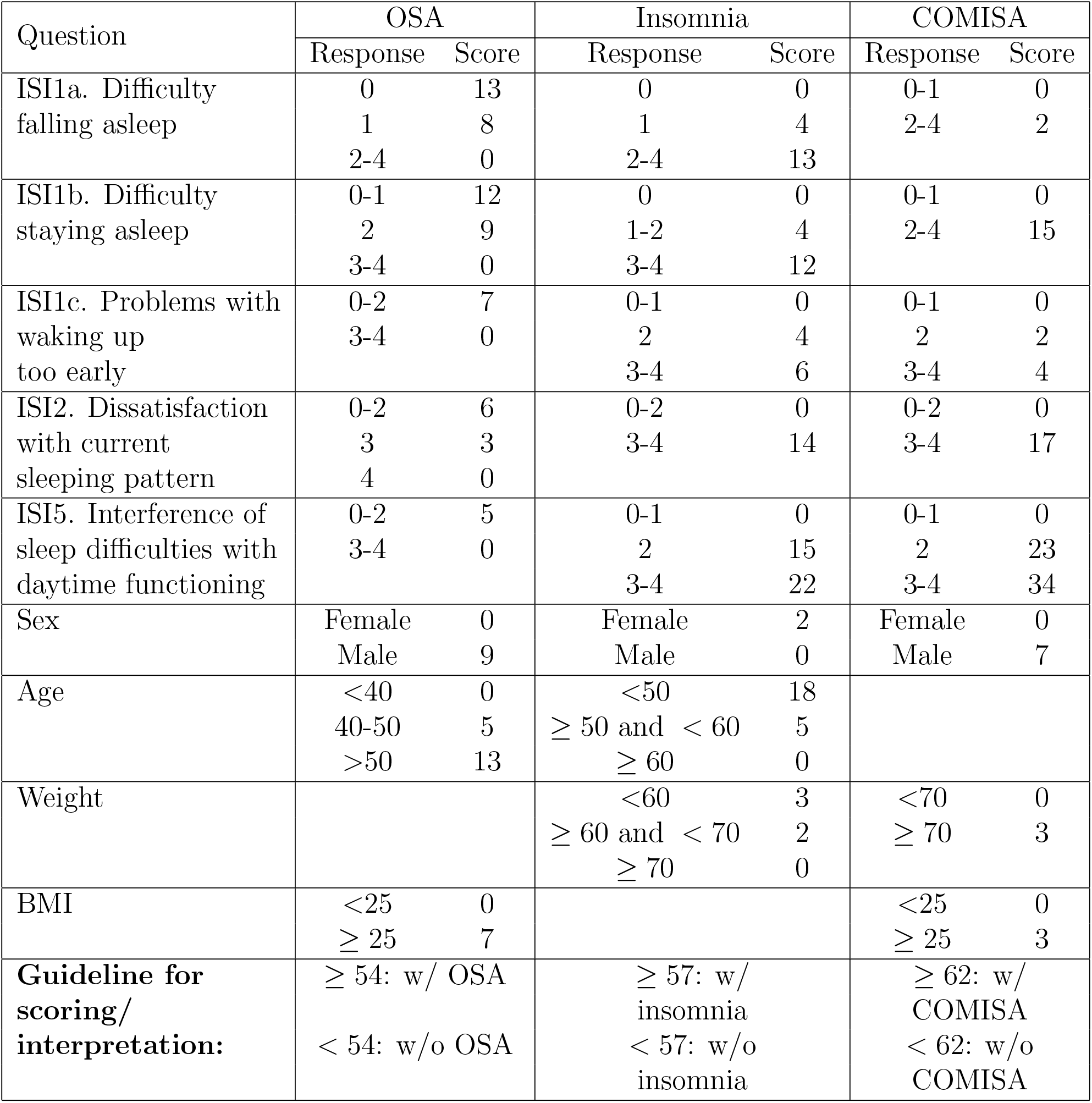
Simplified score table of SLEEPS using SymScore for OSA, Insomnia, and COMISA. Low severity to high severity are represented by the responses 0 to 4 for the Insomnia Severity Index (ISI) items, where 0 indicates no problem and 4 indicates a severe problem. By summing the responses to each question, we can predict the risk of OSA, insomnia, or COMISA. The risk is determined by comparing the total score to the thresholds *Thr*_OSA_ = 54, *Thr*_insomnia_ = 57, and *Thr*_COMISA_ = 62 for OSA, insomnia, and COMISA, respectively, which were obtained from SymScore. If the total score exceeds this threshold, the individual is classified as having the respective sleep disorder.

| Question | OSA |  | Insomnia |  | COMISA |  |
| --- | --- | --- | --- | --- | --- | --- |
|  | Response | Score | Response | Score | Response | Score |
| ISI1a. Difficulty falling asleep | 0 | 13 | 0 | 0 | 0-1 | 0 |
|  | 1 | 8 | 1 | 4 | 2-4 | 2 |
|  | 2-4 | 0 | 2-4 | 13 |  |  |
| ISI1b. Difficulty staying asleep | 0-1 | 12 | 0 | 0 | 0-1 | 0 |
|  | 2 | 9 | 1-2 | 4 | 2-4 | 15 |
|  | 3-4 | 0 | 3-4 | 12 |  |  |
| ISI1c. Problems with waking up too early | 0-2 | 7 | 0-1 | 0 | 0-1 | 0 |
|  | 3-4 | 0 | 2 | 4 | 2 | 2 |
|  |  |  | 3-4 | 6 | 3-4 | 4 |
| ISI2. Dissatisfaction with current sleeping pattern | 0-2 | 6 | 0-2 | 0 | 0-2 | 0 |
|  | 3 | 3 | 3-4 | 14 | 3-4 | 17 |
|  | 4 | 0 |  |  |  |  |
| ISI5. Interference of sleep difficulties with daytime functioning | 0-2 | 5 | 0-1 | 0 | 0-1 | 0 |
|  | 3-4 | 0 | 2 | 15 | 2 | 23 |
|  |  |  | 3-4 | 22 | 3-4 | 34 |
| Sex | Female | 0 | Female | 2 | Female | 0 |
|  | Male | 9 | Male | 0 | Male | 7 |
| Age | <40 | 0 | <50 | 18 |  |  |
| | 40-50 | 5 | $\geq 50$ and $< 60$ | 5 | | |
| | >50 | 13 | $\geq 60$ | 0 | | |
| Weight |  |  | <60 | 3 | <70 | 0 |
| | | | $\geq 60$ and $< 70$ | 2 | $\geq 70$ | 3 |
| | | | $\geq 70$ | 0 | | |
| BMI | <25 | 0 |  |  | <25 | 0 |
| | $\geq 25$ | 7 | | | $\geq 25$ | 3 |
| **Guideline for scoring/ interpretation:** | $\geq 54$: w/ OSA | | $\geq 57$: w/ insomnia | | $\geq 62$: w/ COMISA | |
|  | < 54: w/o OSA |  | < 57: w/o insomnia |  | < 62: w/o COMISA |  |

In terms of specificity, XGBoost outperforms SymScore across all sleep disorders, with higher values for OSA (0.72 vs. 0.62), insomnia (0.85 vs. 0.78), and COMISA (0.87 vs. 0.80). While SymScore sacrifices a small degree of specificity, it compensates with higher or comparable sensitivity, maintaining a balance that is still clinically valuable. These results indicate that SymScore prioritizes correctly identifying true positive cases, which is crucial in clinical settings where under-diagnosis can lead to serious health consequences. While its specificity is lower, the balance achieved by SymScore allows for effective screening, minimizing missed cases while avoiding excessive false positives. Overall, these sensitivity and specificity values highlight SymScore’s utility in clinical practice, supporting its potential for reliable screening and risk assessment in sleep disorder prediction.

We verified that the models were not overfitting by comparing their performances on both the training and testing sets (Table S5). Additionally, using K-fold cross-validation across different data subsets, we confirmed that the models were stable and generalizable.

Regarding the float variables, SymScore also offers significant advantages over AutoScore. That is, unlike AutoScore, SymScore determines optimal response groupings for float variables. For example, SymScore divides BMI into two groups—below 25 (with weight 0) and 25 or above (with weight 7)—to maximize predictive accuracy for OSA (Table 6 column 2). This is different from manual grouping based on quantiles used in AutoScore.

By using Table 6, we can simply get the sum of an individual’s responses. By comparing this sum with the threshold for each disorder, we can classify whether or not an individual has a sleep disorder. The thresholds identified with SymScore are *Thr*_OSA_ = 54 for OSA, *Thr*_insomnia_ = 57 for insomnia, and *Thr*_COMISA_ = 62 for COMISA. If the total sum of a patient’s responses exceeds the respective threshold, the patient is classified as having that particular sleep disorder. This demonstrates the convenience of SymScore questionnaires for clinical applications, offering a straightforward method for disorder classification.

In addition to simplifying classification, SymScore score tables provide valuable insights into the factors influencing these disorders. For instance, for COMISA, questions ISI1b, ISI2, and ISI5 (with weights ranging from 15 to 34) were found to significantly impact classification, compared to other ISI questions (with weights between 2 and 4) as shown in Table 6 column 4. For OSA and insomnia, the weight related to age reveals how the probability of developing each disease changes with age (Table 6 columns 2 and 3). These results illustrate that SymScore generates easily interpretable shortened questionnaires with high performance, making it a practical tool for clinical settings.

## 4. Discussion

In this work, we introduced the Symbolic Regression-Based Clinical Score Generator (SymScore), a novel approach designed to simplify and enhance the accuracy of health risk assessments in clinical settings. Traditional machine learning-based approaches, while effective, are often considered ‘black boxes’, lacking transparency and interpretability. They also require specialized expertise and equipment, making them costly and challenging to implement in real-world adoption. SymScore addresses these challenges by generating simplified, interpretable questionnaires based on symbolic regression, enabling ease of use and a high level of accuracy.

SymScore leverages genetic programming-based symbolic regression to automatically categorize responses to selected questions, streamlining the questionnaire creation process while maintaining predictive accuracy. We validated SymScore’s performance against the MCQI-6 and SLEEPS questionnaires, which are established machine learning-based tools for diagnosing sleep disorders. SymScore’s score tables offer predictive accuracy comparable to that of these tools. Additionally, SymScore’s simplified structure makes it easier to interpret, in contrast with the complexity often associated with machine learning-based approaches.

To generate a score table, users need to simply input an Excel file containing survey response data and then specify the task—either regression, to estimate disease risk based on total score, or classification, to determine disease presence. Additionally, users can input essential details such as the range of responses for the questions and the corresponding maximum values. SymScore then optimally groups responses and assigns weights, reflecting their importance. These response groups and their corresponding weights are then used to generate a score table, allowing users to easily sum responses to assess disease risk. For regression tasks, the sum indicates risk, while for classification, SymScore calculates a threshold to classify disease presence. This transparent and straightforward process enables healthcare professionals to understand and trust the predictions without requiring extensive computational resources or specialized expertise.

The importance of simplicity and transparency in clinical tools is further highlighted by the AutoScore framework developed by Xie et al., which combines machine learning with regression techniques to create point-based scoring models [51]. While AutoScore manually groups responses, SymScore automates this task, enhancing accuracy and efficiency. Moreover, SymScore integrates important clinical constraints like monotonicity in response weights, ensuring that the generated scores accurately reflect disease severity—a critical improvement. Additionally, SymScore supports both classification and regression tasks, offering greater versatility than AutoScore. Unlike the P-ROSC score implementation of AutoScore [60], which converts raw scores into probabilities, SymScore’s scoring system is inherently interpretable, avoiding the need for such conversions.

Another notable interpretable tool is FEAT (Feature Engineering Automation Tool), developed by La Cava et al. [56]. While FEAT also employs symbolic regression, it categorizes input values into only two groups based on a threshold, limiting its ability to capture nuanced relationships between features and disease severity. In contrast, SymScore’s method of optimal response grouping and precise weight assignment, along with monotonicity constraints, effectively captures these complexities. Note that while FEAT has been tested on datasets with increasing feature relationships, its performance on datasets with non-increasing relationships has not been evaluated.

Despite the existence of other interpretable clinical decision-making tools, SymScore distinguishes itself by offering advanced features alongside ease of use. This combination facilitates efficient, accurate, and user-friendly risk assessments, ultimately contributing to improved patient outcomes and streamlined healthcare processes. However, it is important to acknowledge the limitations of our study. First, the datasets utilized were specific to certain clinical contexts. Furthermore, SymScore is designed for medical questionnaires that include numerical rating scales and is not suitable for those involving subjective or qualitative responses. Future studies should test SymScore with a broader range of datasets and clinical settings to validate its generalizability. Next, SymScore relies on the assumption that the relationships between questionnaire responses are independent. This assumption may not hold for all types of questionnaires or clinical conditions. Lastly, while SymScore provides substantial interpretability and simplicity, there may be minor trade-offs in predictive accuracy compared to more complex machine learning models. Further refinements to the algorithm could enhance the balance between accuracy and interpretability.

SymScore represents a significant advancement in explainable AI, offering transparent and interpretable models suitable for clinical practice. By bridging the gap between complex machine learning models and practical clinical applications, SymScore delivers a user-friendly, resource-efficient, and interpretable tool for generating shortened questionnaires. While SymScore has been applied specifically to questionnaires, its flexible and generalizable framework shows potential for broader clinical applications. This versatility underscores SymScore’s contribution to clinical risk assessment, combining ease of use with robust performance and paving the way for wider adoption and improved patient care.

## 5. Conclusion

Our study introduces SymScore, a powerful and user-friendly tool designed to simplify and enhance health risk assessment questionnaires in clinical practice. SymScore addresses the interpretability challenges often associated with traditional machine learning-based questionnaires by providing a transparent, explainable scoring system through the use of symbolic regression. It automatically optimizes response grouping and applies monotonic constraints, ensuring that each score meaningfully reflects disease severity. By offering intuitive score tables with easily interpretable response scores, SymScore eliminates the need for specialized expertise or significant computational resources, making it ideal for real-world clinical use.

SymScore’s performance has been validated against high-accuracy tools, MCQI-6 and SLEEPS, for sleep disorder assessment, achieving comparable predictive accuracy while offering superior interpretability. This balance of accuracy and transparency positions SymScore as a valuable resource in explainable AI, bridging the gap between advanced machine learning and practical, clinician-friendly applications in patient care. By facilitating straight-forward, accurate risk assessments, SymScore has the potential to improve clinical workflows and enhance patient outcomes.

## Data Availability

All data produced in the present study are available upon reasonable request to the authors

## 6. Data Availability

The MCQI and SLEEPS datasets are not publicly available but are available from the corresponding author on reasonable request.

## 7. Acknowledgments

We thank the following organizations for their support of this study: Institute for Basic Science (Institute for Basic Science Grant IBS-R029-C3 to J.K.K), Samsung Medical Center (Samsung Medical Center Grant OTC1190671 to E.Y.J), and Hyundai Motor’s Chung Mong-Koo Global Scholarship (to O.R.C).

