## Supplementary Material for "SymScore: Machine Learning Accuracy Meets Transparency in a Symbolic Regression-Based Clinical Score Generator"

### Contents

S1 Parameter Tuning s2
S2 Correlations between key items in MCQI-6 and SLEEPS questionnaires s4
S3 Details of Optimal Response Grouping s4
S4 Details of Threshold Computation s6
S5 Cross-validation and checking for overfitting s6

*Corresponding author
† These authors contributed equally to this work.

### S1. Parameter Tuning

The performance parameter tuning for the symbolic regression models involves carefully selecting key parameters that influence the genetic programming algorithm's ability to generate effective candidate solutions. To search for optimal parameter values, we employed Bayesian optimization. The parameter descriptions and values used in the parameter tuning process are shown in Table S1.

The function_set only includes the operation addition in order to focus on additive relationships, which is particularly relevant to the nature of the problem at hand, which is assigning weights to key questionnaire items whose sum estimates the actual total score. This also simplifies the search space and allows for faster optimization.

The p_crossover is set at 0.7 to encourage diversity among candidate solutions. By setting p_crossover to 0.7, we prioritize the blending of successful characteristics from high-performing candidate solutions, which is crucial for efficient search in the solution space. This value allows for the maintenance of diversity while ensuring convergence toward optimal solutions.

In contrast, the probabilities for p_subtree_mutation, p_hoist_mutation, and p_point_mutation are allowed to vary within a narrower range of 0 to 0.1. This decision reflects a controlled approach to introducing variations in the candidate solutions. Given that these mutations can significantly alter the structure of solutions, limiting their probabilities helps prevent excessive disruption that may lead to losing promising candidates. The range of 0 to 0.1 allows for experimentation with slight variations, allowing innovation while preserving the strengths of the best candidate solutions.

For generations, values are set at 50, indicating the total number of iterations the algorithm will execute to select the best candidate solutions and apply genetic operations. The population_size is defined as 20000, specifying the number of candidate solutions maintained in each generation. The stopping_criteria parameter is set to 0.01 for regression tasks with mean absolute error (MAE) as the corresponding fitness metric and $10^{-4}$ for classification tasks with the area under the receiver operating curve (AUROC) as the fitness metric. This determines when the algorithm should terminate based on the fitness metric, indicating minimal improvements in performance.

Finally, the max_samples parameter is set at 0.9 to ensure that the model is trained on a substantial portion of the data for better generalization.

| Parameters | Description | Values |
| --- | --- | --- |
| `function_set` | Defines which operations the genetic programming algorithm can use to construct candidate solutions | ‘add’ |
| `p_crossover` | The probability of applying crossover, which involves combining parts of two parent solutions to produce new offspring candidate solutions | 0.7 |
| `p_subtree_mutation` | The probability of applying subtree mutation, which involves replacing a randomly chosen subtree in a candidate solution with a new randomly generated subtree | $0 \sim 0.1$ |
| `p_hoist_mutation` | The probability of applying hoist mutation, which involves simplifying a candidate solution by replacing a subtree with one of its subtrees that is smaller in size | $0 \sim 0.1$ |
| `p_point_mutation` | The probability of applying point mutation, where a random node in an expression tree is changed (e.g., changing an operator) | $0 \sim 0.1$ |
| `generations` | The total number of iterations the algorithm will run. Each generation consists of selecting the best candidate solutions and applying genetic operations (crossover, mutation) | 50 |
| `population_size` | Defines the number of candidate solutions in each generation | 20000 |
| `stopping_criteria` | Parameter that determines when the algorithm should stop based on the fitness metric chosen | 0.01 (regression)<br>$10^{-4}$ (classification) |
| `max_samples` | Parameter that specifies the maximum fraction of the training samples to be used to fit each candidate solution in the population | 0.9 |

Table S1: The parameter values used for tuning the symbolic regression model

The integration of these parameters into the optimization process, supported by Bayesian techniques, allows for a systematic exploration of the parameter space, ultimately leading to improved performance and robustness in the training of the symbolic regression model.

### S2. Correlations between key items in MCQI-6 and SLEEPS questionnaires

| | Q23 | Q28 | Q39 | Q51 | Q58 | Q60 |
|---|---|---|---|---|---|---|
| Q23 | 1.000 | 0.579 | 0.371 | 0.507 | 0.398 | 0.506 |
| Q28 | 0.579 | 1.000 | 0.321 | 0.612 | 0.429 | 0.589 |
| Q39 | 0.371 | 0.321 | 1.000 | 0.420 | 0.408 | 0.350 |
| Q51 | 0.507 | 0.612 | 0.420 | 1.000 | 0.448 | 0.590 |
| Q58 | 0.398 | 0.429 | 0.408 | 0.448 | 1.000 | 0.557 |
| Q60 | 0.506 | 0.589 | 0.350 | 0.590 | 0.557 | 1.000 |

Table S2: Correlation table for key items in the MCQI-6 shortened questionnaire

| | ISI1a | ISI1b | ISI1c | ISI2 | ISI5 | age | weight | BMI | sex |
|---|---|---|---|---|---|---|---|---|---|
| ISI1a | 1.000 | 0.588 | 0.429 | 0.497 | 0.451 | 0.130 | -0.177 | -0.083 | 0.256 |
| ISI1b | 0.588 | 1.000 | 0.659 | 0.581 | 0.490 | 0.231 | -0.131 | -0.053 | 0.158 |
| ISI1c | 0.429 | 0.659 | 1.000 | 0.461 | 0.399 | 0.253 | -0.100 | -0.038 | 0.095 |
| ISI2 | 0.497 | 0.581 | 0.461 | 1.000 | 0.644 | 0.058 | -0.040 | -0.001 | 0.113 |
| ISI5 | 0.451 | 0.490 | 0.399 | 0.644 | 1.000 | 0.079 | -0.048 | 0.003 | 0.115 |
| age | 0.130 | 0.231 | 0.253 | 0.058 | 0.079 | 1.000 | -0.247 | -0.086 | 0.103 |
| weight | -0.177 | -0.131 | -0.100 | -0.040 | -0.048 | -0.247 | 1.000 | 0.869 | -0.502 |
| BMI | -0.083 | -0.053 | -0.038 | -0.001 | 0.003 | -0.086 | 0.869 | 1.000 | -0.227 |
| sex | 0.256 | 0.158 | 0.095 | 0.113 | 0.115 | 0.103 | -0.502 | -0.227 | 1.000 |

Table S3: Correlation table for key items in the SLEEPS shortened questionnaire

### S3. Details of Optimal Response Grouping

We use step functions to group the answers of each input feature. Because we utilize symbolic regression (GPlearn python package), the step functions are automatically generated as part of the regression process, resulting in automatic grouping. In GPlearn, symbolic regression is based on genetic programming, where the population for the new generation is created through a series of genetic operations such as selection, crossover, mutation, and reproduction, applied to individuals in the current population. After these operations, the individuals for the next generation are selected based on their fitness, which is typically calculated by their error values (how well they fit the data). GPlearn uses symbolic regression to search for mathematical expressions that fit the data, so the computational time and memory usage scale with the size of the data. Symbolic regression also includes various operators (e.g., addition, multiplication, trigonometric functions). The more operators we include in the search space, the larger the search space becomes, increasing the computational cost. Therefore, the computational complexity can be expressed as $O(g \times p \times s \times m \times n)$, where

$g$: the number of generations
$p$: the population size (number of candidate expressions per generation)
$s$: the number of allowed operators
$m$: the complexity of the expressions
$n$: the size of the dataset

### S4. Details of Threshold Computation

The default threshold probability is set to 0.5 in Eq. 7, given that the expression output is a probability-like value. Specifically, we generate score tables where each possible response is assigned a value, and the total sum of these scores is proportional to the severity of a specific disease. To convert this total sum into a probability, we apply the sigmoid function, $\sigma(x) = \frac{1}{1+e^{-x}}$. When the total sum is input into the sigmoid function, a bias term b is added. This bias is determined through the genetic programming process in GPlearn, as part of the evolved symbolic expression. Additionally, the parametrization of the sigmoid function can affect the ROC curve. A steeper or shifted sigmoid curve could result in different probability distributions for the predicted classes. In our model, we used the standard sigmoid function without any additional parameterization, as we found it sufficient for balancing sensitivity and specificity.

### S5. Cross-validation and checking for overfitting

| MAE on training set | MAE on test set | Cross-validated MAE | Mean cross-validated MAE | SD of cross-validated MAE |
|---|---|---|---|---|
| 9.182 | 10.381 | 10.718 | 10.718 | 1.084 |

Table S4: **Evaluation metrics for checking for overfitting in the SymScore model for the MCQI-6 questionnaire**

| | AUROC on training set | AUROC on test set | Cross-validated AUROC | Mean cross-validated AUROC | SD of cross-validated AUROC |
|---|---|---|---|---|---|
| OSA | 0.859 | 0.843 | 0.850 | 0.850 | 0.007 |
| insomnia | 0.905 | 0.910 | 0.895 | 0.898 | 0.025 |
| COMISA | 0.904 | 0.932 | 0.903 | 0.906 | 0.004 |

Table S5: **Evaluation metrics for checking for overfitting in the SymScore model for the SLEEPS questionnaire**

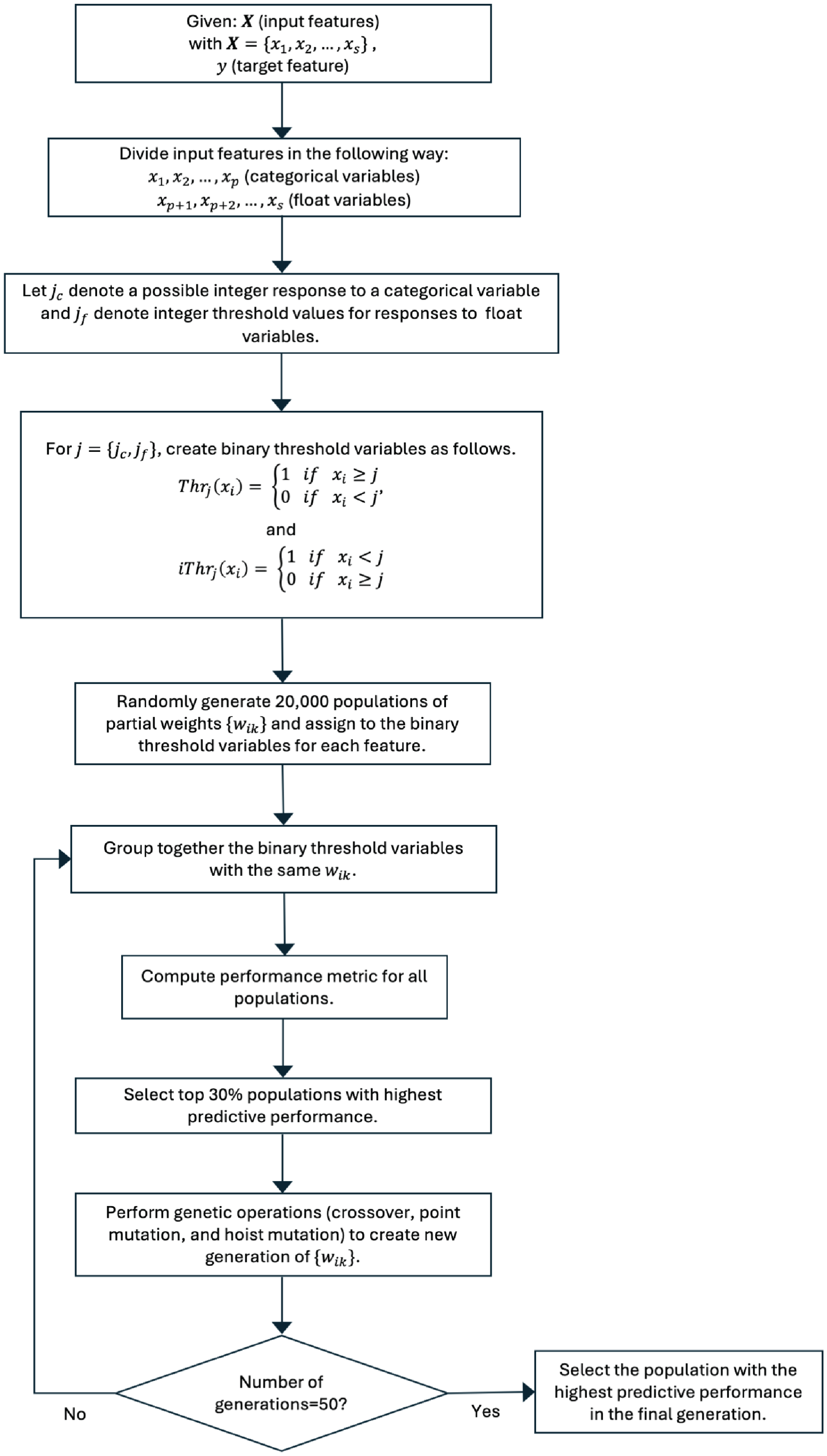


Figure S1: **Diagram illustrating the optimal response grouping process.**
